# Correlation between the environmental parameters with outbreak pattern of COVID-19: A district level investigation based on yearlong period in India

**DOI:** 10.1101/2021.06.28.21259631

**Authors:** Amitesh Gupta, Laboni Saha

## Abstract

The present study has investigated the role of regional meteorology and air quality parameters in the outbreak pattern of COVID-19 pandemic in India. Using the remote sensing based dataset of 12 environmental variables we correlated infective case counts at a district level in India. Our investigation carried out on the circumstantial data from more than 300 major affected districts in India and found that air quality parameters are playing very crucial role in this outbreak. Among the air pollutants, O_3_ was better correlating with infection counts followed by AOD, CO, NO_2_, BC and SO_2_. We also observed that among the weather parameters air temperature, incoming shortwave radiation, wind speed are positively and significantly associate with outbreak pattern and precipitation and humidity are negatively correlated with confirmed cases; only cloud cover has no significant relation. We noted that coastal districts in the both coast of India and districts located in the plain and low-lying areas have experienced bitter situation during this pandemic. Our study suggests that improving air quality with proper strict regulations and complete lockdown during the peak of pandemic could reduce the misfortune in all over India.

## Introduction

In the 21st century, the world has been battle-scarred by the pernicious ordeal of coronavirus disease (COVID-19) since early 2020. By the end of March, 2021, a total of 152,006,071 confirmed cases with a death toll of 3,193,315 have been registered all over the world (https://www.worldometers.info/coronavirus/). This pneumonia infection caused by Severe Acute Respiratory Syndrome CoronaVirus-2 (SARS-CoV-2) had initially broken out in Wuhan, Hubei province, China during late 2019 (Guan et al., 2020; Liu et al., 2020a; Wu and McGoogan, 2020; Zhu et al., 2020a; Zu et al., 2020) and gradually spread all over the world (Bertuzzo et al., 2020; Gorbalenya et al., 2020). Contemplating the hasty unfurl of this virus and its human-to-human transmissibility from contagion (Wang et al., 2020a; Wang et al., 2020b; Wang et al., 2020c), the World Health Organization (WHO) declared this disease to be a global pandemic on March 11, 2020. India, the domicile of 17.7% of the total world population had noted its very first COVID-19 confirmed case on January 27, 2020, and afterwards a total of 12,220,717 cases have been confirmed by the end of March, 2021 with a death count of 162,960 (https://www.covid19india.org/).

Worldwide sundry clinical inspections of COVID-19 have diagnosed respiratory droplets and fomites from contaminated persons as the spawn agent of this lethal disease (Alag, 2020; Ge et al., 2013; Huang et al., 2020a; Jana et al., 2020; Jung et al., 2021; Kunz et al., 2020; Sathian et al., 2020; Tuttle, 2020). Umpteen researchers also reported about analogous symptoms of this disease to MERS and SARS (Holshue et al., 2020; Perlman, 2020; Tan et al., 2005; Wang et al., 2020c). Since the SARS-CoV-2 virus initially causes respiratory damage and this disease has very close similarities to the influenza in terms of various symptoms like Fever, Cytokine storm syndrome, diffuse alveolar damage, Lymphopenia, Coagulation, and immunocompromised conditions (Harapan et al., 2020; Khorramdelazad et al., 2021; Larsen et al., 2020; Li et al., 2021), a few researchers have already exhorted about significant association of environmental parameters to COVID-19 transmission (Azuma et al., 2020; Bherwani et al., 2020; Eslami and Jalili, 2020; Gupta and Pradhan, 2020a; Kifer et al., 2021; Rume and Islam, 2020; Saadat et al., 2020; Shakil et al., 2020). Mention Worthy, earlier shedload of studies have conjectured about conspicuous impact of weather conditions on dissemination of respiratory viruses and its seasonality (D’Amato et al., 2018; Pica and Bouvier, 2012; Price et al., 2019). There are also a few global as well as regional studies that had been carried out in the context of China, USA, England, Germany, Spain, Italy, Indonesia, Iran, Pakistan, Bangladesh, India, and found significant correlation between weather parameters and the COVID-19 cases (Ahmadi et al., 2020; Chen et al., 2021; Fadli et al., 2020; Ficetola and Rubolini, 2021; Gupta et al., 2020a; Gupta and Pradhan, 2020b; Islam et al., 2020; Liu et al., 2020b; Ma et al., 2020; Mehmood et al., 2021; Nottmeyer and Sera, 2021; Oliveiros et al., 2020; Qi et al., 2020; Tosepu et al., 2020). On a contrary, a few studies have also reported that meteorological observations are not significantly associated with the outbreak pattern (Iqbal et al., 2020; Jahangiri et al., 2020; Jamil et al., 2020; Mollalo et al., 2020; Shi et al., 2020; Xie and Zhu, 2020). Studies carried out by Bochenek et al., 2021; Borah et al., 2020; Briz-Redón and Serrano-Aroca, 2020; Emediegwu, 2021; Guo et al., 2021; Gupta et al., 2020b; Mecenas et al., 2020; Runkle et al., 2020; Şahin, 2020; Sil and Kumar, 2020 suggested that warm and humid condition seem to foreshorten the proliferation of COVID-19. On the other hand, (Al-Rousan and Al-Najjar, 2020; Awasthi et al., 2020; Bashir et al., 2020a; Fawad et al., 2021; Gupta and Pradhan, 2020a; Kumar and Kumar, 2020; Pani et al., 2020; Sangkham et al., 2021; Selcuk et al., 2021; Sharma et al., 2021) reported that hotter days might be more susceptible for COVID-19 dissemination while the role of humidity was still incongruous (Aidoo et al., 2021; Auler et al., 2020; Gupta et al., 2020c; Kumar, 2020; Yuan et al., 2021). Menebo, 2020 found negative association between precipitation and COVID-19 cases, contrary to Byass, 2020; while Gunthe et al., 2020 found no significance of precipitation and cloudiness for this outbreak.

Studies have enunciated that frequent short-term exposure to air pollutants could aggrandize the likelihood of symptomatic acute respiratory infections among adults (Kim et al., 2018; Kirwa et al., 2021; Xing et al., 2016). Chauhan and Johnston, 2003; Zhang et al., 2019a had reported that moderate and long-term exposure to imprudent volume of the oxidant air pollutants like Ozone (O_3_), Nitrogen di-Oxide (NO_2_) have the potential to exacerbate the inflammatory effects of the viral and bacterial respiratory infections. Insufflating such air with prohibitive concentration of Particulate Matters (PM) and Carbon Monoxide (CO) have been also inculpated for escalating the risk of cardiovascular and respiratory diseases, and contributing to the mortality rate inclusively (Huang et al., 2018; Khaniabadi et al., 2017; Liu et al., 2018; Yang et al., 2020a). Recent studies have stated that both short-term and long-term exposure to different air pollutants including PM (PM_2.5_, PM_10_), Sulphur di-Oxide (SO_2_), CO, O_3_ and NO_2_ could contribute significantly to the higher rates of COVID-19 infections in different parts of the world (Ali and Islam, 2020; Bourdrel et al., 2021; Coker et al., 2020; Gujral and Sinha, 2021; Jain et al., 2021; Li et al., 2020; Lin et al., 2020; Ma et al., 2021; Matthew et al., 2021; Mele et al., 2021; Sharma et al., 2021; Suhaimi et al., 2020; Wu et al., 2020), therefore positively associated with the conveyance of COVID-19. O_3_ is also a salient precursor of hydroxyl radical (OH), thus it also entices the concentration of SO_2_ and CO (Logan et al., 1981; Thompson, 1992). On the other hand, the carbonaceous pollutants i.e. CO and BC and OC, which also includes the primary and secondary organic carbon, are mainly produced during different anthropogenic activities of biomass burning and fossil fuel combustion, with relatively little contribution from various natural sources (Li et al., 2016; Prabhu et al., 2020; Schmidt, 2011; Wang et al., 2019; Winiger et al., 2016; Zhang and Wang, 2011). Though the source mechanism of these carbonaceous agents are presumably analogous, its spatial concentration differs due to the complex physio-chemical transformation in wayward meteorology (Saliba et al., 2010; Vautard et al., 2007). AOD, an illustrious indicant of Particulate Matter pollution (Chudnovsky et al., 2014; Engel-Cox et al., 2004; Gupta et al., 2021; Karimian et al., 2016), accounts columnar concentration of gaseous and liquid airborne particles has not been often related to such malady study. Since the significant relation between prevailing weather conditions and regional pollution level is well demonstrated by various studies, it becomes very crucial to behold the role of meteorology and air quality togetherly to conceive the seasonality of such savage seizure of coronavirus.

Scanty studies have been carried out on the importance of freshen air for the melioration of human health and to elude any convalescence (Cragg et al., 2016; Kampa and Castanas, 2008; Manisalidis et al., 2020; White et al., 2019). Likewise, scarcely any study has been fanatical about the significance of weather parameters on COVID-19 recovery, even though 93.88% of the sufferers have been healed in India, slightly lesser than the global recovery rate (97.42%) till March 31, 2021. Since a handful of studies had reckoned that the inferior air quality had efficaciously swayed to aggravate the COVID-19 emanation, hence it becomes also requisite to investigate whether the convivial air quality has been benevolent to convalesce the COVID-19 patients in India. Till date, seldom study might have inspected the yearlong period of pandemic situation while minimal have rendered any estimation on this. Therefore, the present study has been firmed to analyse the association of COVID-19 incidences (i.e. transmission) and recovery cases with weather and air quality parameters across the country, as well as proffered a comparative assessment for estimating confirm and recovery cases availing the non-linear complex relationship.

## Data and Methodology

Counts of Confirmed and Recovered cases for all the available districts in India have been acquired from https://www.covid19india.org/. The 2nd wave of COVID-19 in India has prominently surged April, 2021 onwards, hence we have obtained our study for the period throughout the 1st wave of COVID-19 only. During March, 2020 these counts were too craggy, therefore we acquired data only for the period of April, 2020 to March, 2021. Spatial data for total six different meteorological parameters - 2-m Air Temperature (AT), Bias-corrected Total Precipitation (PRC), Specific Humidity (Hm), Cloud Cover (CLD), Incoming Short-wave Radiation (ISWR), Wind Speed (WS), and six different air quality parameters - Aerosol Optical Depth (AOD), Tropospheric Column Nitrogen di-Oxide (NO_2_), Total Column Ozone (O_3_), Surface concentration of Carbon Monoxide (CO), Sulphur di-Oxide (SO_2_) and Black Carbon (BC) have been incorporated in the present study. Remote Sensing based datasets are highly useful for various environmental studies (Bhatt et al., 2021; Das et al., 2017; Das and Gupta, 2021; Gupta et al., 2020b; Gupta et al., 2019; Moniruzzam et al., 2018; Nanda et al., 2018; Rousta et al., 2020), hence we have used such dataset from different sources. Monthly mean of these twelve environmental variables were further processed in ArcGIS software, adjoined and related with the monthly cumulative counts of confirmed and recovery cases for each months for each of those selected districts.

### Infections and recoveries during 1st COVID-19 waves in India

The district wise spatial distribution of COVID-19 confirmed cases and recoveries during the 1st COVID-19 wave (up to March, 2021) are shown in Figure 1. It shows that 35.03% districts had noted cases within the range of 10,000 to 1,00,000; while only 3.05% districts in India registered more than 1 lakh cases during this study period. Noteworthy, most of the districts have registered counts of infective cases within the range of 2500-5000 following the range of 10000-25000. Comprehensively, 21 states registered more than 1 lakh infected cases; among them only Maharashtra and Kerala had registered more than 10 lakh cases. The spatial distribution of cases also depict that the coastal districts were more anguished by misfortune than the districts located in the interior part of the country whereas the districts from the Himalayan Mountains and its foothills had lesser strife during this pandemic. The temporal pattern of daily cases of infections, recoveries, deceased, and test positivity (%) from all over the country were shown in Figure 2a. It depicts that the peak of pandemic was during the early September, 2020 when the daily death counts and confirmed cases were highest, however daily test positivity was highest during July. These trends were at their trough during the winter followed by the post-monsoon seasons but again started to incline since March, 2021.

**Figure 1.**
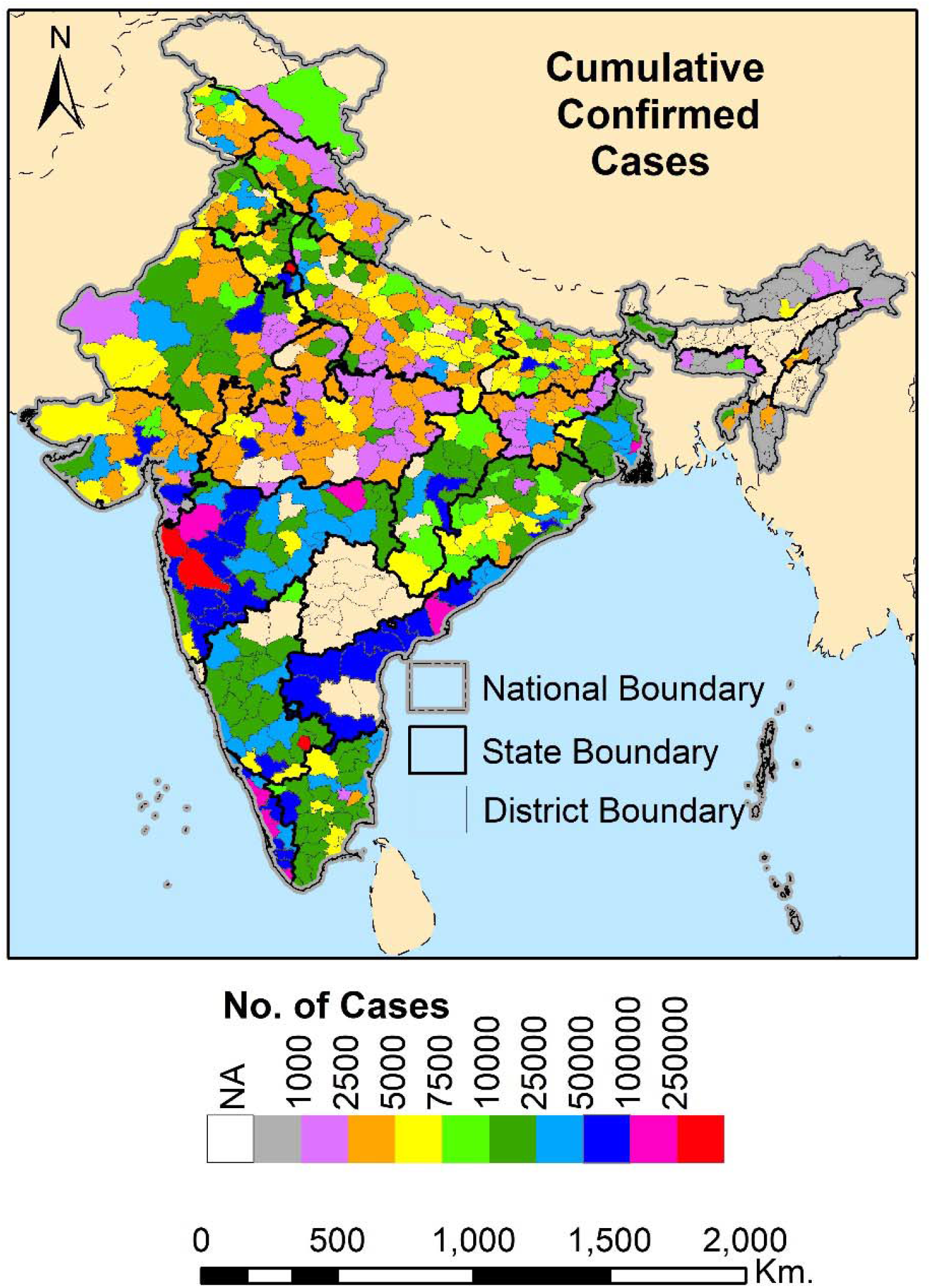
Distribution of confirmed cases

**Figure 2.**
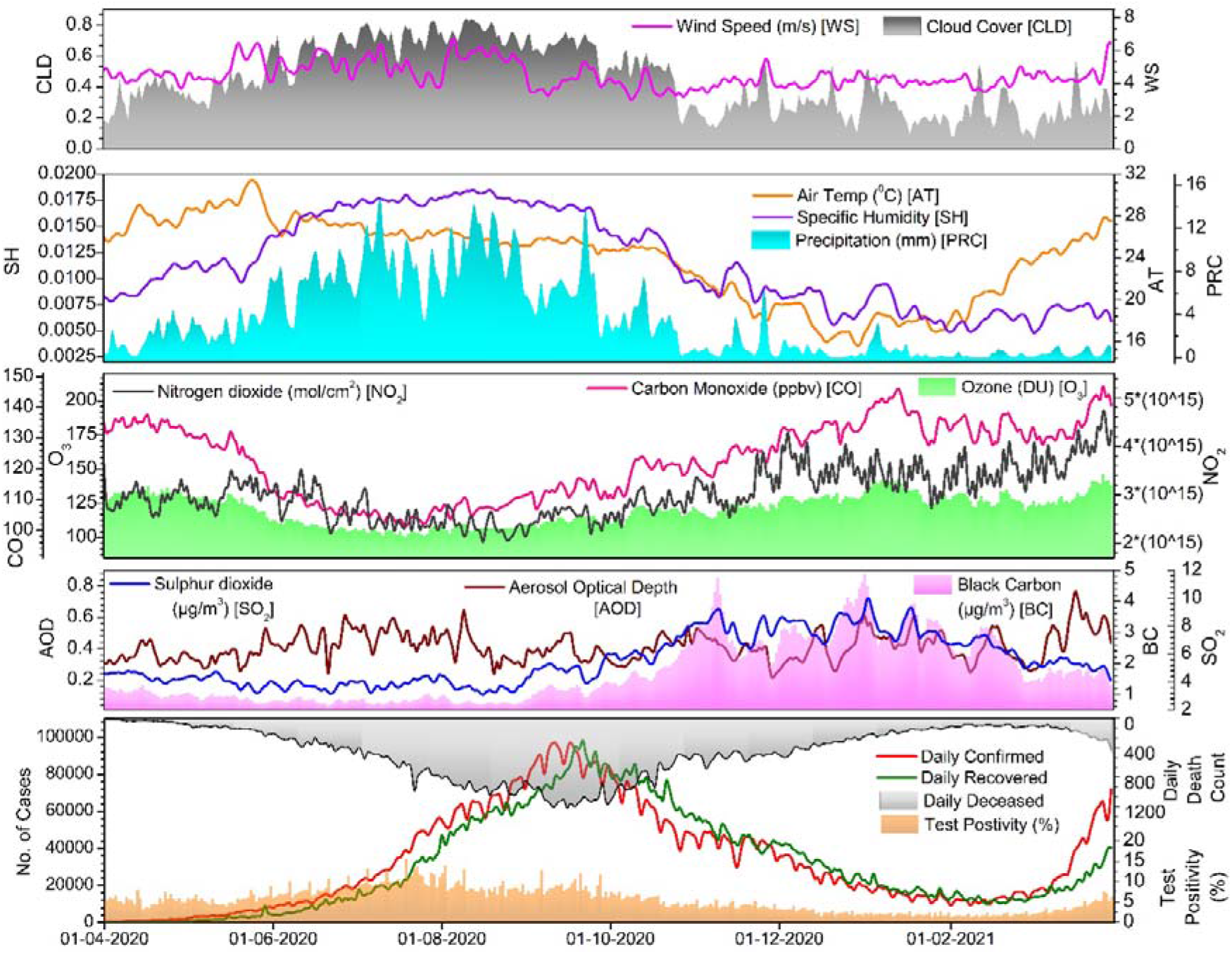
temporal trends

### Pearson’s correlation test

Pearson’s correlation technique was applied to explore the degree of association of environmental parameters with infected and recovered cases at 95% confidence interval using equation 1. A correlogram is also being prepared to better represent the interrelation of the variables of the input dataset.

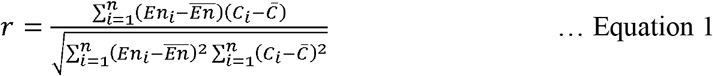

Here, *r* is the correlation coefficient, *En* represents the environmental variables, *C* denotes the count of confirm and recovery cases individually.

## Results and Discussion

Pearson’s correlation test (Figure 3) found that confirmed cases have significantly (p<0.05) positive correlation with the AT (0.483), ISWR (0.268) and WS (0.391), negative association with Hm (−0.304) and PRC (−0.288); however positive but non-significant (p>0.05) relation with CLD (0.109). It depicts that areas with areas with comparatively higher AT with greater ISWR and drier atmospheric conditions (lower humidity and lesser precipitation) seem to register more infected cases in India, quite similar to what was observed by Aslam et al., 2020 and Islam et al., 2020 over Pakistan and Bangladesh respectively. Since the Indian landmass is located in the tropical latitudes of the Northern Hemisphere where the regional ISWR is higher than the global average ISWR during April to September (Hatzianastassiou et al., 2005), thus avails to put up the air temperature during these summer months. Thus, it may indirectly help viruses to spread out at higher temperature conditions. On the other hand, Dowell, 2001; Lowen et al., 2007 had shown that the transmission of respiratory virus via aerosol could be suppressed by higher humid conditions, but enhanced under dry conditions. Barreca, 2012; Dalziel et al., 2018 also suggested that low-humidity levels could be considered as an imperative risk factor for respiratory infection diseases (Tellier, 2006). Thus, the summer conditions become more susceptible for this viral transmission in India. A few studies such as Coccia, 2021; Rendana, 2020; Rosario et al., 2020 have opined that higher wind could lower the emanation of the virus, however we found persuasive correlation between wind and COVID-19 cases in India, thus agreeing to Babu et al., 2020; Gautam et al., 2021. Another crucial reason behind this might be that the districts located near and along the western coast in Kerala, Maharashtra as well as the districts of Tamilnadu, Andhra Pradesh and Odisha located in the eastern coast of India catalogued maximum cases during May to July, 2020 which is the onset duration of monsoon over Indian subcontinent, thus experience higher wind from the sea-side towards the land. Liu et al., 2020c had suggested that the rapid weather variability might result in an increase in respiratory infections in warming climatic conditions; moreover, study by Moriyama and Ichinohe, 2019 had found that higher ambient temperature could restrain the adaptive immune response to a virus infection which also could be accorded through the present study.

**Figure 3.**
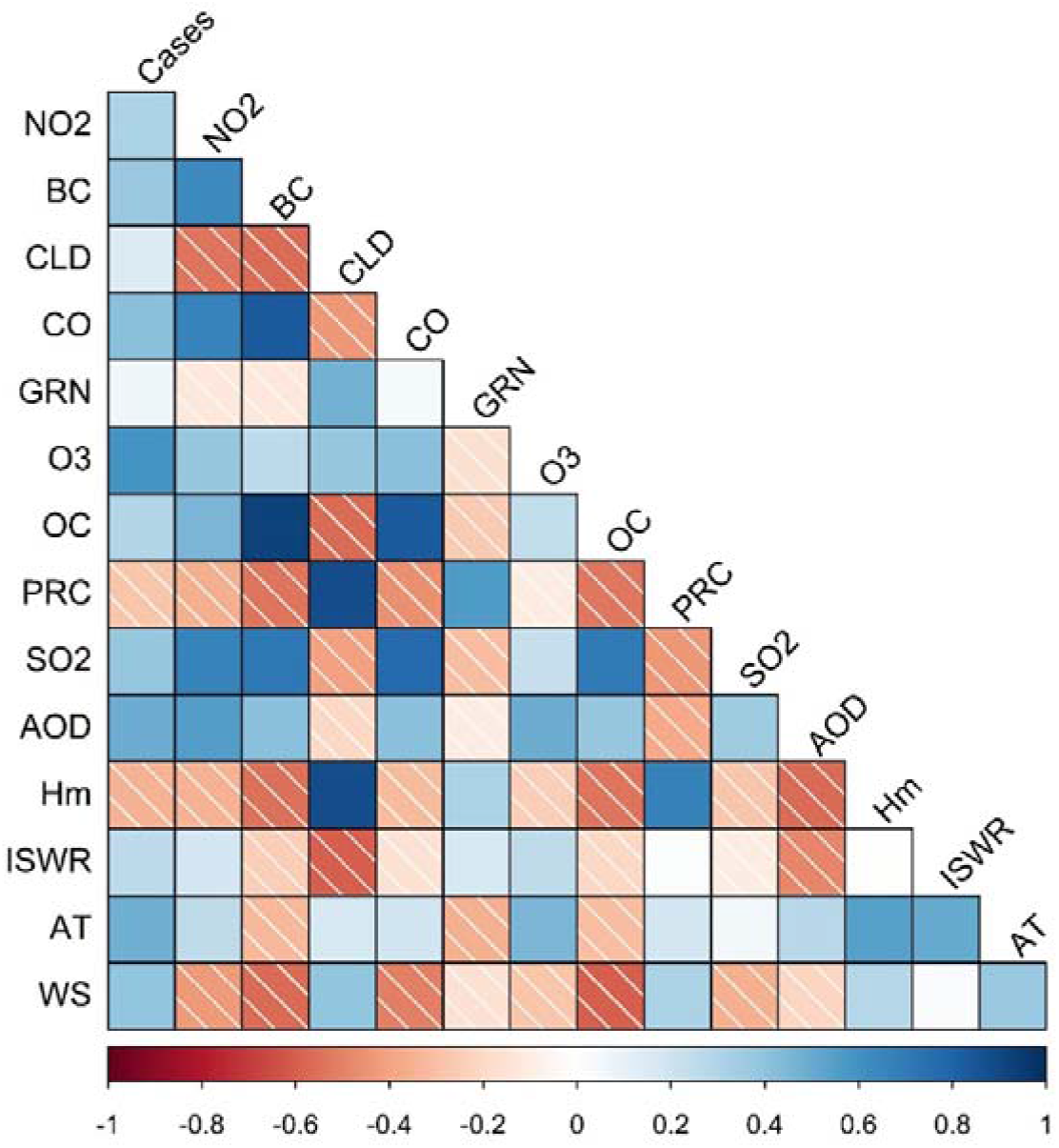
correlogram

In agreement of numerous scientific research conducted in the last two decades, the link between environmental pollution and chronic illnesses mainly the cardiovascular and respiratory diseases are quite comprehensible (Andersson et al., 2020; Chauhan and Johnston, 2003; Ghude et al., 2016; Mehta et al., 2013; Orru et al., 2017; Zhang et al., 2019b; Ciencewicki and Jaspers, 2007). In the current study we found all the seven air quality parameters to be significantly and positively correlated with confirmed cases. Figure 4 depicts that monthly confirmed cases was better correlated with monthly average concentration of O_3_ (0.595) followed by AOD (0.497), CO (0.414), NO_2_ (0.383), BC (0.374) and SO_2_ (0.325). It confers that concentration of oxidant pollutants had a significant impact on COVID-19 transmission in Indian scenario. Earlier studies had conjectured the O_3_ as a mighty oxidant (Nuvolone et al., 2018; Turner et al., 2015; Zhang et al., 2019c). Likewise, exposure to NO_2_ has been found to be associated with the increasing risk of acute respiratory tract infections (Munawer, 2018; Suryadhi et al., 2020; Valko et al., 2007). On the other hand, CO is thoroughly soluble in the bloodstream (Power, 1968). It becomes toxic when blood reacts with haemoglobin (Badman and Jaffé, 1996; Schraufnagel et al., 2019). There were several hotspots observed in the northern IGP and eastern India contributing detestable amounts of CO and NO_2_ that could be held accountable for the appalling air quality in those regions as it might have significantly swayed the exposed individuals to get intensely infected by the coronavirus. Therefore, higher concentration of O_3_ along with its precursors (CO and NO_2_) could be held noxious for such awful viral emanation especially over northern and eastern part of India. Thus we agreed with Bashir et al., 2020; Li et al., 2020; Pansini and Fornacca, 2020; Zhu et al., 2020b; Zoran et al., 2020 about the significance of O_3_, with CO and NO_2_ pollution regarding the COVID-19 emanation. Furthermore, in developing countries like India, the organic fuel based power generation and daily household combustion activities are commonly observed which often results in excessive emission of SO_2_ and carbonaceous pollutants such as CO and BC. A beguiling study by Hahon et al., 1985 with a rigorous snooping on mice had found that the severity of influenza virus infection was better recognizable with a prominent exposure to diesel engine emissions and coal dusts. In the present study, we have found a rich correlation between pollutants emitted due to fossil fuel combustion and COVID-19 outbreak patterns. Till date, a minimal study has obtained SO_2_ and BC to investigate the surging of COVID-19. The pathophysiological corroboration cogently conveyed pathways of various diseases arose from SO_2_ and NO_2_ pollution (Brunekreef and Holgate, 2002; Chen et al., 2015; Kampa and Castanas, 2008). The elevated concentration of SO_2_ can shorten the lung function, causing suffocation, wheezing, coughing, and also arouse the exasperation in the tissues inside the nose and the throat (Feuyit et al., 2019) (Chen et al., 2007; Kelsall et al., 1997). It was quite distinctive that higher SO_2_ and coronavirus outbreak were experienced over IGP, Odisha, coastal areas of Gujarat and Maharashtra. The spatial distribution of BC also depicts an approximate pattern of higher concentration over IGP and eastern coast of Odisha. In various parts of the world, these pollutants have been adjudged as an indicator of adverse health effects (Janssen et al., 2011; Segersson et al., 2017; Tobias et al., 2014). It becomes virulent (Bourdrel et al., 2017; Kampa and Castanas, 2008), thus damaging the lung functions (Jacobson et al., 2014; Paulin and Hansel, 2016; Paunescu et al., 2019; Valavanidis et al., 2008; Xu et al., 2018a). Therefore, it can be quite distinctive that the ensued elevated pollution due to outrageous fossil fuel consumption could cause the neighbourhood to be less immune and predisposed to severe respiratory infection like COVID-19 (Kyung and Jeong, 2020; Suhaimi and Jalaludin, 2015; Thurston et al., 2017), hence prominently associated with pulmonary diseases (Anenberg Susan C. et al., 2010; Peters et al., 2001; Rosenthal et al., 2013). Since PM concentration can’t be directly assessed through Satellite based observations (Gupta et al., 2021, 2006; Gupta and Christopher, 2009; Li et al., 2015), we correlated satellite derived AOD with infective cases over several districts during the study period, and found that the IGP was limned with exorbitant AOD where the coronavirus outbreak was also quite austere. Earlier, Spekreijse et al., 2013; Zhao et al., 2019 recommended the strong possibilities of airborne transmission of respiratory virus via fine dust particles. Recently, Comunian et al., 2020; Tang et al., 2020 also discussed the evidence of SARS-CoV-2 virus transmission via dust particles. Therefore, considering the elevated AOD over semi-arid north-west India along with scorching summer conditions, and the infective cases from Rajasthan, Punjab, Haryana and Uttar Pradesh during April-June. Interaction between the meteorological parameters and air pollutants’ concentrations are quite complex and varying seasonally, and often influenced by the local physiography including the topographic variations such as location of mountains, valleys, or the presence of substantial water bodies such as large rivers, and lakes (Benjamin, 1975; Eum et al., 2015; Hewson and Olsson, 1967; Mahapatra et al., 2019; Prabhu et al., 2020; Saikawa et al., 2019; Zhang et al., 2018). During summer, the rising AT and comparatively lesser SH ominously abet the pollutants’ concentration by physio-chemical transformation (Analitis et al., 2018; Jayamurugan et al., 2013; Sario et al., 2013), henceforth make warmer days commendatory for such virus infections. While comparing with the long term average conditions, many studies had acknowledged the imposition of four tier countrywide lockdown for relatively cleaner air in India during April to mid of June, 2020 (Gupta et al., 2020d; Naqvi et al., 2021; Nigam et al., 2021; Singh and Chauhan, 2020; Venter et al., 2020). Even though we found that the IGP followed by the coastal districts of West Bengal, Odisha, Andhra Pradesh, Maharashtra, Kerala, Tamilnadu had inhaled quite bad air where each states had registered more than 30,000 cases during that period. Intriguingly, the temporal trends of pollutants showed comparatively higher concentration during winter and post-monsoon seasons, while the infections were relatively lesser in both those seasons. Therefore, it suggests that the shoddy air quality could not be discretely blameworthy for coronavirus dissemination in India, rather bad air with successively increasing AT in summer environment could be held responsible.

## Conclusion

Our conscientious study suggests that the summer tropical environment in India may spare a more affirmative condition for SARS-CoV-2 transmission, agreeing with Sajadi et al., 2020; Xie and Zhu, 2020. Instead of squashing the COVID-19 curve, higher air temperature may significantly assist an augment in pandemic predicament in India. Comparatively higher wind in drier areas could indulge the virus to unfurl more, as also observed by Aidoo et al., 2021; Babu et al., 2020. Besides, the negative correlation between precipitation and infections, also discerned by Mansouri Daneshvar et al., 2021, illustrates the relevance of prevailing atmospheric conditions in dissemination of this pandemic. Scientific communities’ acquiescence on the adverse impact of air quality on crude mortality is inevitable in any well-being circumstances across the world (Bentayeb et al., 2015; Davidson et al., 2005). Foreseeably, the poor air quality was perceived to be significantly correlated with the COVID-19 dissemination during the study period. The positive correlation between coal blazed pollution and COVID-19 outbreak, as well as the negative association between the recovery count from this disease and prevailing regional pollution levels stoutly suggest that the exorbitant air pollution due to bounteous non-renewable energy generation and its perpetual consumption might perturb the inbred immunity of residents in circumjacent area, and hinder them also in agile recovery from this disease. The concurrent heath tribulation and the interconnection of diseases affecting multiple organs are also decisively discerned during COVID-19 infections. During or in a later stage of treatment, several patients are also condemned to various post-syndrome infections mostly related to cardiovascular and respiratory disorders which might be the ramifications of virulent vulnerability to the gaseous pollution. Hence it altogether asserts that the environment needs to be ameliorated immensely by its standards to alleviate such deadly pandemic. The injurious impact of traffic pollution has also been discussed through various global and regional studies (Gauderman et al., 2007; Laumbach and Kipen, 2012; Urman et al., 2014). The pre-eminence of weather parameters on air quality is indubitable and evinced globally (Bhaskar and Mehta, 2010; Fisher, 2002; Yansui Liu et al., 2020e; Panofsky and Prasad, 1967; Qin et al., 2020; Seo et al., 2018; Zhang et al., 2015), hence its significant association with COVID-19 dissemination during a year-long period over India has been certainly revealed through the present study.

## Data Availability

This study has used open source data. COVID-19 data has been collected from https://www.covid19india.org and remote sensing based data were collected from GIOVANNI.

## Notes

### Competing Interest Statement

The authors have declared no competing interest.

### Funding Statement

No fund has been provided to carry out this study.

### Author Declarations

RBased Services Pvt. Ltd.

